# Large language models for accurate disease detection in electronic health records

**DOI:** 10.1101/2024.07.27.24311106

**Authors:** Nils Bürgisser, Etienne Chalot, Samia Mehouachi, Clement P. Buclin, Kim Lauper, Delphine S. Courvoisier, Denis Mongin

**Affiliations:** Division of Rheumatology, Department of Medicine, Geneva University Hospitals, Geneva, Switzerland; Division of General Internal Medicine, Department of Medicine, Geneva University Hospitals, Geneva, Switzerland; Quality of Care Division, Geneva University Hospitals, Geneva, Switzerland; Faculty of medicine, University of Geneva, Geneva, Switzerland; Information systems directorate, Geneva University Hospitals, Geneva, Switzerland

## Abstract

**Importance:** The use of large language models (LLMs) in medicine is increasing, with potential applications in electronic health records (EHR) to create patient cohorts or identify patients who meet clinical trial recruitment criteria. However, significant barriers remain, including the extensive computer resources required, lack of performance evaluation, and challenges in implementation.

**Objective:** This study aims to propose and test a framework to detect disease diagnosis using a recent light LLM on French-language EHR documents. Specifically, it focuses on detecting gout (“goutte” in French), a ubiquitous French term that have multiple meanings beyond the disease. The study will compare the performance of the LLM-based framework with traditional natural language processing techniques and test its dependence on the parameter used.

**Design:** The framework was developed using a training and testing set of 700 paragraphs assessing “gout”, issued from a random selection of retrospective EHR documents. All paragraphs were manually reviewed and classified by two health-care professionals (HCP) into disease (true gout) and non-disease (gold standard). The LLM’s accuracy was tested using few-shot and chain-of-thought prompting and compared to a regular expression (regex)-based method, focusing on the effects of model parameters and prompt structure. The framework was further validated on 600 paragraphs assessing “Calcium Pyrophosphate Deposition Disease (CPPD)”.

**Setting:** The documents were sampled from the electronic health-records of a tertiary university hospital in Geneva, Switzerland.

**Participants:** Adults over 18 years of age.

**Exposure:** Meta’s Llama 3 8B LLM or traditional method, against a gold standard.

**Main Outcomes and Measures:** Positive and negative predictive value, as well as accuracy of tested models.

**Results:** The LLM-based algorithm outperformed the regex method, achieving a 92.7% [88.7-95.4%] positive predictive value, a 96.6% [94.6-97.8%] negative predictive value, and an accuracy of 95.4% [93.6-96.7%] for gout. In the validation set on CPPD, accuracy was 94.1% [90.2-97.6%]. The LLM framework performed well over a wide range of parameter values.

**Conclusions and Relevance:** LLMs were able to accurately detect disease diagnoses from EHRs, even in non-English languages. They could facilitate creating large disease registries in any language, improving disease care assessment and patient recruitment for clinical trials.

**Key points:** *Question:* How accurate and efficient are large language models (LLMs) in detecting diseases from unstructured electronic health records (EHR) text compared to traditional natural language processing techniques?

*Findings:* This study proposes a framework based on Meta’s Llama 3 8B, a recent public LLM, outperforming traditional natural language processing techniques in detecting gout and calcium pyrophosphate deposition disease in unstructured text. It achieves high positive and negative predictive values and accuracy. Performance was robust over a wide range of parameters.

*Meaning:* The proposed framework can ease the use of LLMs in effectively detecting disease in EHR data for various clinical applications.

## Introduction

Since the release of their first version in 2018 ^1^, Large Language Models (LLM) have improved very quickly, with frequent application in research ^2^. They are effective for many complex task in the medical field such as passing advanced exams ^3^, answering patients questions ^4^ or interpreting and extracting clinical concepts and data ^5,6^. They could thus be of high interest to interpret, categorize or extract information from Electronic Health Records (EHR), especially from the unstructured text (i.e. free-text) of the hospital documents, as recently proven for the classification of injuries ^7^.

In a previous work, to build a self-updating gout registry from hospital EHR data ^8^, we used regular expressions (regex) and natural language processing techniques to identify patients with gout from any hospital documents. The task proved to be rather complex: in addition to situations in which the diagnosis is negated or aiributed to a family member, the word “goutte” (gout in French) also designs drops and droplets, which are common words used to designate quantity of drugs or body fluids. It took months of back-and-forth to specify the query algorithm able to avoid unexpected uses of the word “gout”. LLM, on the other hand, because of their advanced natural language understanding abilities ^9^, could be able to correctly identify diagnosis with little additional work.

The aim of this article is to propose a general framework for using LLM to detect specific disease diagnoses with the goal of creating automatic EHR registries or facilitating patient recruitment for clinical trials. We test the ability of advanced LLM such as those of the Llama family ^10^, using few shot prompting and chain of thought techniques ^11^, to identify patients with a gout diagnosis from unstructured EHR data of a tertiary university hospital. The performance of the procedure is assessed by comparing the prediction to a gold standard and evaluating it against the performance of a regular expression (regex)-based algorithm. We perform extended sensibility analysis to assess the robustness of our results against LLM parameter change or against prompt structure ^12^. We further validate the proposed LLM framework on the detection of another form of crystal arthropathy, calcium pyrophosphate deposition disease (CPPD), from EHR documents. Ease of implementation and computing resources needed are also considered.

## Methods

### Objective

We aim to test and propose a framework for the LLM allowing to detect a disease diagnosis in an unstructured EHR document. We used a challenging disease to detect in French, gout (“goutte”), to evaluate the framework’s effectiveness, as “goutte” can also mean drops or droplets, be used for proverb or even surname, in other contexts.

First, The LLM and a regex-based algorithm were used to determine if the sentence containing the word “goutte” indicates a positive diagnosis, or something else (negative diagnosis, diagnosis of a family member, other use of the word “goutte”). In a second step, the LLM framework was validated on a second dataset to determine if sentences containing words referring to calcium pyrophosphate deposition disease (i.e. chondrocalcinosis, pseudogout) indicate a positive diagnosis of CPPD or not. CPPD, a subform of calcium pyrophosphate deposition disease, is a frequent differential diagnosis of gout.

### Setting

The data stemmed from electronic documents of the Geneva University Hospital (HUG), a 2’000 beds tertiary hospital and Swiss’ largest, serving a population of 517,802 residents as well as neighboring French Nationals working in Switzerland. A random sample of 1000 documents containing the word “goutte” (gout) and 500 containing the CPPD-related words “chondrocalcinose” (chondrocalcinosis) or “pyrophosphate” or “pseudogoutte”(pseudogout) were queried from the data-lake of the HUG, a mongoDB mirrored and centralised database version of the EHR data. From these documents, the surrounding of the detected words (“goutte” for gout, and CPPD-related terms for CPPD) were extracted, considering a 30 words boundary before and after the detected word, or sentence punctuation, whichever is closest. These short paragraphs were then manually anonymised before being analysed by the two algorithms. The resulting sentences were then split in three datasets:

- A training one concerned with gout, used to tune our algorithms.
- A testing one concerned with gout, used to estimate their performances.
- A validation one, concerned with CPPD, used as a validation for the proposed LLM framework.

### Gold standard

The selected phrases were evaluated by two health-care professionals (one internal medicine physician, one registered nurse, both trained by a board-certified rheumatologist) to assess if they described the patient as having the disease gout for the training and testing dataset and CPPD for the validation dataset, opposed to a negation of the diagnosis, the description of family diagnosis (e.g. the father had gout), or an alternate use of the word (“goutte” for drugs, body fluids or other uses). Disagreements were resolved by a board-certified rheumatologist.

### Regex algorithm

The regex algorithm to detect gout diagnosis consisted in the following steps:

1. Normalization of the text (removing special characters, accents, duplicated space, setting to lower case)
2. Extracting the context of each “goutte” word, the context being considered as 8 words before and 5 after the word “goutte”.
3. Performing the following tests for each resulting context:
  - Presence of a drug that can be administrated in droplets (based on the list of all drugs allowed in Switzerland);
  - Presence of a human body liquid, as an indication that “goutte” refers to the droplet;
  - Presence of a French expression using the word “goutte”;
  - Presence of the words “family”, “father”, “mother”, “sister”, “brother” as an indication that the diagnosis concern a family member;
  - Presence of a negative word and no double negation, as an indication for a negative diagnosis.
4. Combining the tests. The patient is considered as having gout, if all words in the context are not detected by the tests in step 3.

Details and code can be found at: https://gitlab.unige.ch/goutte/register_validation

### Few shots prompting of Llama 3

We used an 8-bit quantized version of the Meta-Llama-3-8B-Instruct model. Meta-Llama-3-8B is the smaller model of the Meta Llama 3 family of large language. The instruct version has been tuned and optimized for dialogue use case. The 8-bit quantized version allowed us to use easily accessible GPU, such as NVIDIA Titan X with 12 GB of VRam, in conjunction with 20G of RAM standard CPU.

The prompt (entire prompt can be found at https://gitlab.unige.ch/goutte/llm_detection_of_diagnosis) consisted in the following steps:

1. The description of the role of the algorithm (“you are a text classifier aiming at identifying gout diagnosis”)
2. A paragraph of context (What is gout, in what context can the word “goutte” be used)
3. Description of the 2 categories considered (positive gout diagnosis, or other)
4. The structure of the expected response with a simple chain of thought structure ^13^:
  - A short explanation detailing the reasoning.
  - The result category, based on the short explanation, after the string “A:”.
5. A set of 10 examples with the desired output (technique called few shots prompting). The output of the LLM was then parsed following the expected structure.

The parameter set used was a temperature of 0.3, a repetition penalty of 1, and cumulative probability and most likely next word sampling (top_p and top_k), with top_k = 40 and top_p = 0.95.

Sensitivity analysis: The effect of the temperature and the penalty parameters were tested by varying the temperature from 0.1 to 0.8, and the penalty parameter from 0.8 to 1.4. The impact of the prompt was tested by iteratively removing the steps described previously, or by increasing the number of result categories to 4 (positive gout diagnosis, negative gout diagnosis, liquid, or other). Stability of the answer for the standard parameter set was assessed by performing 5 successive classifications for each sentence.

### Statistics

We summarised data using frequencies and percentages for categorical variables and median and interquartile range for continuous variables. Positive predictive value (PPV) was calculated as the proportion of documents referring to the disease gout correctly detected by the model among all documents referring to the disease. Negative predictive value (NPV) was calculated as the proportion of documents not referring to the disease correctly detected by the model among all non-disease documents. Accuracy was defined by the sum of correct outcome according to the gold standard, divided by the total number of documents tested.

Confidence intervals were computed using the Wilson method ^14^. All statistics were computed using the software R 4.2.0 ^15^.

### Ethical consideration

The use of the gout registry^8^ data for quality improvement programs has been approved by the Geneva ethics commission (CCER 2023-00129). The need for consent was waived by the Geneva Ethics Committee because this study qualifies as a quality improvement initiative

## Results

Of the documents in the testing dataset, we analyzed 757 sentences containing the word “goutte” in French, of which 235 sentences (31%) indicated that the patient had the disease gout (according to the manually reviewed charts, see table 1). Of the documents in the validation dataset, we analyzed 600 sentences, of which 376 (62%) indicated a positive diagnosis of CPPD.

**Table 1:**
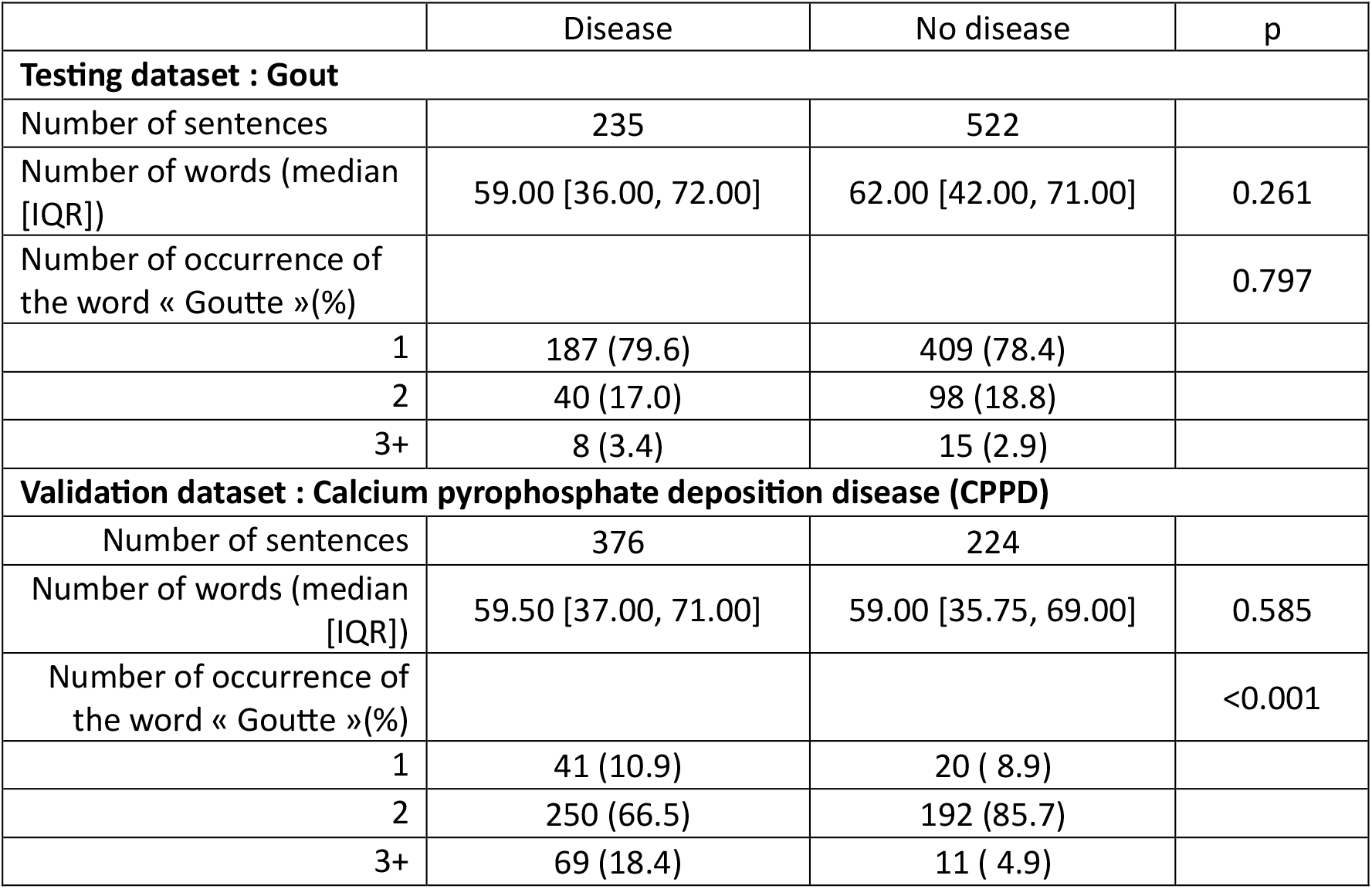
characteristics of the testing and validation dataset.

|  | Disease | No disease | p |
| --- | --- | --- | --- |
| **Testing dataset : Gout** |  |  |  |
| Number of sentences | 235 | 522 |  |
| Number of words (median [IQR]) | 59.00 [36.00, 72.00] | 62.00 [42.00, 71.00] | 0.261 |
| Number of occurrence of the word « Goutte »(%) |  |  | 0.797 |
| 1 | 187 (79.6) | 409 (78.4) |  |
| 2 | 40 (17.0) | 98 (18.8) |  |
| 3+ | 8 (3.4) | 15 (2.9) |  |
| **Validation dataset : Calcium pyrophosphate deposition disease (CPPD)** |  |  |  |
| Number of sentences | 376 | 224 |  |
| Number of words (median [IQR]) | 59.50 [37.00, 71.00] | 59.00 [35.75, 69.00] | 0.585 |
| Number of occurrence of the word « Goutte »(%) |  |  | <0.001 |
| 1 | 41 (10.9) | 20 ( 8.9) |  |
| 2 | 250 (66.5) | 192 (85.7) |  |
| 3+ | 69 (18.4) | 11 ( 4.9) |  |

### Algorithm performance

The LLM based algorithm tended to perform better than the regex based one (figure 1), reaching similar positive predicting value (PPV) (92.7% [88.7-95.4%] compared to 92.3% [87.9-95.2%]) but a slightly higher negative predicting value (NPV) (96.6% [94.6-97.8%] compared to 92.3% [89.8-94.3%]) The LLM algorithm had an overall accuracy of 95.4% [93.6-96.7%] slightly higher than the 92.3% [90.2-94.0%] accuracy of the regex-based algorithm. Accuracy of both the regex-based and the LLM algorithms in the calibration sample were slightly higher (97.8% [96-98.7%] and 96.8% [95.5-98.4%] respectively for the Llama3 and regex-based algorithm) than in the validation set.

**Figure 1:**
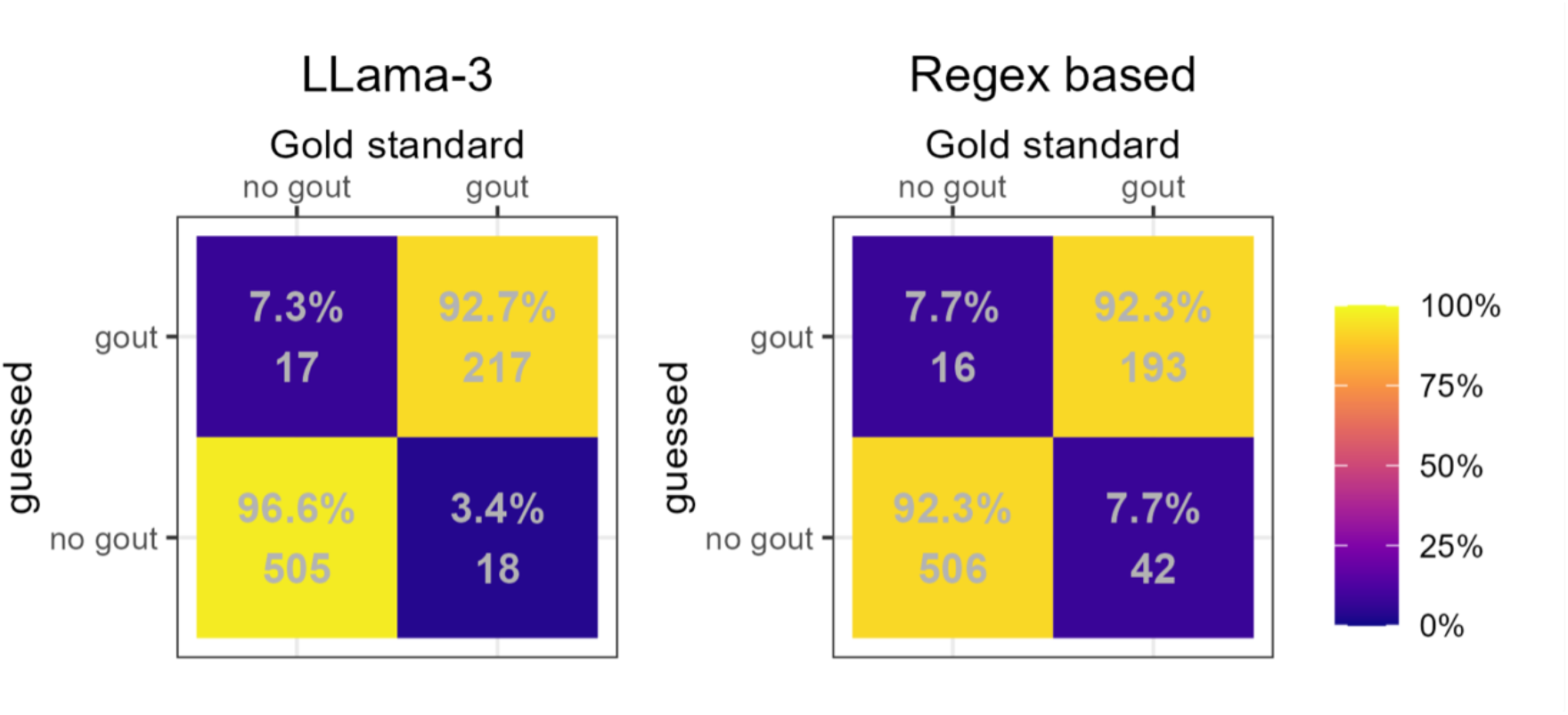
confusion matrix of the two algorithms, compared to the gold standard.

The regex-based algorithm took a few seconds to run on the 757 sentences on a standard laptop, while the llama3 algorithm took one hour and a half using a CPU with 20GB of RAM and a 12GB VRAM GPU.

### Sensitivity analysis

The results of the LLM based algorithm were robust against parameter changes: similar performance was obtained for temperature ranging from 0.1 to 0.8, and for penalty parameter from 0.8 to 1.3. Penalty parameter of 1.4 resulted in the LLM tending to produce non structured output, which could not be parsed (see supplementary eTable 1 and 2).

Concerning the effect of the prompt (table 2):

**Table 2:** Sensitivity analysis of Llama 3 model with varying prompts.

| Prompt type | Outputs without result | Predictive positive value | Negative predictive value | Accuracy |
| --- | --- | --- | --- | --- |
| Reference (few shot with context, chain of thoughts and 2 output categories) | 0 | 92.7% [88.7-95.4%] | 96.6% [94.6-97.8%] | 95.4% [93.6-96.7%] |
| few shot with context, chain of thoughts and 4 categories output | 0 | 90.2% [85.8-93.3%] | 97.3% [95.5-98.4%] | 95% [93.2-96.3%] |
| few shot with context, no Chain of thoughts and 4 output categories | 0 | 87% [82.2-90.6%] | 95.9% [93.8-97.3%] | 93% [91-94.6%] |
| One shot with context, chain of thoughts and 4 output categories | 0 | 81.5% [76.5-85.7%] | 97.9% [96.2-98.9%] | 91.9% [89.8-93.7%] |
| Zero shot with context, chain of thoughts and 4 output categories | 13 | 84.6% [79.7-88.6%] | 97.6% [95.8-98.6%] | 93.2% [91.1-94.8%] |
| Zero shot with no context, chain of thoughts and 4 output categories | 65 | 63.5% [58.3-68.4%] | 99.4% [97.9-99.8%] | 81.5% [78.4-84.2%] |
| zero shot with context, no Chain of thoughts and 4 output categories | 0 | 89% [83.9-92.6%] | 89.8% [87-92%] | 89.6% [87.2-91.5%] |

- Adding classifying categories did not result in better prediction and even lowered slightly the PPV (90.2% [85.8-93.3%] with 4 categories).
- A one-shot approach (only one example) or a zero-shot approach (without examples) produced lower accuracy, mainly due to lower predictive positive value. The one-shot prompts yielded indeed a PPV of 81.5% [76.5-85.7%] and the zero-shot prompts a PPV of 84.6% [79.7-88.6%]. Of note, zero shot prompts produced several outputs that did not respect the formatting expected, resulting in 65 unusable outputs.
- For the zero-shot prompt:.
  - Removing the context strongly lowered the PPV from 84.6% [79.7-88.6%] PPV to 63.5% [58.3-68.4%]
  - Removing the chain of thought, although allowing more robust output format, resulted in a lower accuracy: from 93.2% [91.1-94.8%] to 89.6% [87.2-91.5%]

When testing 5 inferences for each sentence, the result proved to be the same for the 5 inferences in 98% of the cases. The classification of 8 sentences changed in one of the five inferences, and the classification of 11 sentences changed in two of the five inferences.

### Validation

Reusing the LLM with the same parameters and the same prompt structure on a different disease (CPPD) yielded a PPV of 92.3% [88.4-95.5%] for detecting the presence of CPPD diagnosis, an NPV of 95.9% [91.4-98.1%], for an overall accuracy of 94.1% [90.2-97.6%].

## Discussion

In this study, Llama 3, a recent Large Language Model (LLM), showed excellent positive and negative predictive values in identifying gout diagnosis from unstructured (i.e. free-text) medical documents of electronic health records (EHR) of a French-speaking tertiary university hospital. Performance was slightly better than a regex-based algorithm. The tested prompt structure appears to be a promising template for accurately detecting specific diseases, facilitating the creation of fast and easy-to-implement registries.

Previous studies using natural language technique with or without machine learning have been made to detect gout flare, but prior to or without the use of new LLM ^16,17^. A recent study, using a protected health information compliant form of GPT-4 which extracted hepatological imaging report data from an EHR, showed a similar accuracy ^6^, while the use of LLM to classify injuries showed close to perfect classification capabilities ^7^.

The technique based on regular expressions performed well but required iterative adaptations of the different tests to appropriately reject false positive outcome. For such technique, the transposition to another disease will be language and context dependent, and thus remain time-consuming. Although the LLM technique also needed some iterative work to adapt the prompt and the temperature parameter to obtain proper outputs, the sensitivity study showed that it is robust over a large range of parameter values. Our validation using the proposed framework to detect another disease diagnosis confirms its versatility. It may allow easy implementation in other language or for another condition without expensive tuning. The fact that the LLM algorithm had a great performance using only a two-category output, that is without the need to describe the different situation that could lead to false positive, is clearly an advantage. The slightly lower PPV obtained for the validation dataset when compared to the testing one can be explained by the fact that CPPD in our EHR documents was frequently evocated in differential diagnosis lists, a situation for which the gold standard did not consider it as a diagnosis, but the LLM model tended to do so.

The computing power needed for the LLM model was higher than the regex-based method, which needed only a personal computer. This emphasizes the need for hospitals to provide secure computing power to researchers and clinicians to foster research in free-text documents.

Nevertheless, our study suggests that lighter models of LLM, which do not require extensive computing resources, may also be of significant interest for such applications.

There are some limitations to our study. First, our methods were tested in a single academic institution, though it covers all medicine specialties. Second, the study was conducted using only one language (French). Llama LLM is known to perform well in English, French, German and Spanish ^18^, but results may change strongly for lower resource languages. Secondly, at this stage, we only identified diagnosis, without the identification of specific situation (i.e. gout flare, chronic gouty arthritis, etc.).

## Conclusion

LLM can accurately detect disease diagnosis in EHR’s clinical documents, even when the disease name has a high consonance with other words. Our study proposes a framework that can be easily reused and suggests that LLM can perform well even in language outside their primary training dataset, paving the way for detecting disease with minimal effort, from the rich EHR clinical notes and documents of a hospital. Combined with appropriate resources from hospitals, the template proposed in this study could significantly accelerate the creation of registries and the detection of patients for clinical trial recruitment, reducing the typically months-long process to just days.

## Data Availability

All prompts and code have been made available at the following gitlab repository: https://gitlab.unige.ch/goutte/llm_detection_of_diagnosis. Due to medical confidentiality, we are unable to share the sentences and document data. However, if authorization is obtained from the ethics committee, we may be able to provide access to the data

## Acknowledgment

Authors’ contributions: Designed research: DM, DSC. Performed research: DM, NB, DSC. Analyzed data: DM and EC. Establishment of the Gold Standard: NB, SM, KL, CB. Wrote the paper: NB, DM and DSC. Critical revision of the paper: all authors. DM had full access to all the data in the study and takes responsibility for the integrity of the data and the accuracy of the data analysis

The authors have no conflict of interest to declare.

We thank Emmanuel Durand at the Information Systems Directorate for his help in accessing the database of the Geneva University Hospitals, as well as support for the implementation of the large language model.

## Funding

This project was funded by the Private Foundation of the Geneva University Hospitals, a not-for-profit foundation.

## Notes

### Competing Interest Statement

The authors have declared no competing interest.

### Author Declarations

This study involves human participants and the creation and use of the register for quality improvement programs has been approved by the Geneva ethics commission (CCER 2023-00129). The need for consent was waived by the Geneva Ethics Committee because this study qualifies as a quality improvement initiative.

