## Supplementary material for "Large language models for accurate disease detection in electronic health records": Suplement table 1 and 2

eTable 1: sensitivity study when varying the temperature of the Llama3 model.

| Temperature | Without result | Predictive positive value | Negative predictive value | accuracy |
|---|---|---|---|---|
| 0.1 | 0 | 92.7% [88.7-95.4%] | 96.6% [94.6-97.8%] | 95.4% [93.6-96.7%] |
| 0.2 | 0 | 93.1% [89.1-95.7%] | 96.6% [94.6-97.8%] | 95.5% [93.8-96.8%] |
| 0.3 | 0 | 92.3% [88.2-95.1%] | 96.6% [94.6-97.8%] | 95.2% [93.5-96.5%] |
| 0.4 | 1 | 92.8% [88.7-95.4%] | 96.7% [94.8-98%] | 95.5% [93.8-96.8%] |
| 0.5 | 0 | 93.5% [89.5-96%] | 96.2% [94.2-97.5%] | 95.4% [93.6-96.7%] |
| 0.6 | 0 | 92.4% [88.3-95.1%] | 96.9% [95.1-98.1%] | 95.5% [93.8-96.8%] |
| 0.7 | 0 | 94.3% [90.5-96.6%] | 96.2% [94.2-97.5%] | 95.6% [93.9-96.9%] |

eTable 2: sensitivity study when varying the penalty parameter of the Llama3 model.

| Penalty parameter | Without result | Predictive positive value | Negative predictive value | accuracy |
|---|---|---|---|---|
| 0.7 | 6 | 92.8% [88.8-95.5%] | 96.9% [95-98.1%] | 95.6% [93.9-96.9%] |
| 0.8 | 6 | 93.2% [89.2-95.7%] | 96.9% [95-98.1%] | 95.7% [94-97%] |
| 0.9 | 1 | 92.8% [88.7-95.4%] | 96.7% [94.8-98%] | 95.5% [93.8-96.8%] |
| 1.0 | 0 | 92.7% [88.7-95.4%] | 96.6% [94.6-97.8%] | 95.4% [93.6-96.7%] |
| 1.1 | 0 | 91.7% [87.5-94.5%] | 97.1% [95.3-98.2%] | 95.4% [93.6-96.7%] |
| 1.2 | 0 | 93.6% [89.7-96.1%] | 96.9% [95.1-98.1%] | 95.9% [94.2-97.1%] |
| 1.3 | 0 | 92.3% [88.2-95.1%] | 96.4% [94.4-97.7%] | 95.1% [93.3-96.4%] |
| 1.4 | 13 | 93.6% [89.5-96.1%] | 94.9% [92.6-96.4%] | 94.5% [92.6-95.9%] |
